# The impacts of viral interaction on household transmission of respiratory viruses

**DOI:** 10.1101/2024.07.19.24310723

**Authors:** Jessica C. Ibiebele, Elie-Tino Godonou, Amy P. Callear, Matthew R. Smith, Rachel Truscon, Emileigh Johnson, Marisa C. Eisenberg, Adam S. Lauring, Arnold S. Monto, Sarah Cobey, Emily T. Martin

## Abstract

The possibility of virus-virus interaction has been hypothesized at the population level, but there are limited data regarding the effects of viral interaction on transmission. This study used data from a prospective household cohort study to examine how viral interaction impacts transmission of influenza A virus (IAV), influenza B virus (IBV), and respiratory syncytial virus (RSV). We used two main predictors of transmission: coinfection in the index case and detection of a virus other than a primary virus of interest in the susceptible contact. Household- and individual-level analyses were conducted for IAV, IBV, and RSV. To estimate the risk of transmission within households with coinfected index cases, a household-level analysis was performed using multivariable regression with Poisson mixed effects models. To estimate an individual’s risk of transmission when multiple viruses co-circulated within their household, an individual-level analysis was performed using mixed-effects logistic regression models. Coinfection among index cases was associated with reduced transmission of IAV and RSV. Infection with a different virus among household contacts was associated with an increased risk of transmission of IAV and RSV. This study enhances the understanding of viral interaction by elucidating the potential impacts of interaction on virus transmission. These findings could have important implications for public health planning and prevention efforts.

## Introduction

Acute respiratory infections (ARIs) are a major cause of morbidity and mortality worldwide.^1,2^ In the United States, influenza viruses and respiratory syncytial virus (RSV) contribute to substantial societal and economic burden, especially impacting young children, adults aged 65+ years, and individuals with underlying conditions.^3–5^ Understanding factors that impact transmission of these respiratory viruses is crucial for estimating population risk and developing public health prevention strategies.

One factor that may impact respiratory virus transmission is the interaction between multiple viruses when both infect a single host. The possibility of virus-virus interaction, a phenomenon where circulation of one virus impacts the level of a second virus, has long been speculated based on population-level patterns. Theoretically, viruses may interact with each other positively, exhibiting synergism, or negatively/antagonistically.^6^ The exact mechanisms of interaction that may exist at the individual level are not fully understood, nor are the implications of virus-virus interaction for transmission. Specifically, there is a paucity of data regarding viral transmission in the presence of more than one virus within a household—for example, whether viruses are more or less likely to transmit when an index case is coinfected or when a susceptible contact is infected with a different virus. Notably, some viruses have been shown to have different viral loads when coinfecting with other viruses compared to infecting on their own.^7,8^ Whether such impacts on viral load through viral interference affect transmission is unclear.

Here we examine the possible impacts of viral interaction on transmission of influenza A virus (IAV), influenza B virus (IBV), and RSV within households. We used data from the Household Influenza Vaccine Evaluation (HIVE) study—a prospective cohort study in Michigan—from years 2010-2020 to assess the relationship between the co-circulation of multiple viruses within a household and transmission to susceptible household members.

## Methods

### Study population

The HIVE study is an ongoing prospective cohort study of households with children in Southeast Michigan, U.S.A. that began in 2010 and allows for the study of multiple respiratory pathogens and their transmission dynamics within households. The present study utilized data from HIVE study years 2010-2020. Additional details about the study population have been previously described.^9^ Written informed consent (paper or electronic) was obtained from adults (aged >18). Parents or legal guardians of minor children provided written informed consent on behalf of their children. Participants were compensated for their time and effort. The HIVE study is approved by the institutional review board at the University of Michigan Medical School (HUM00034377 & HUM00118900).

### Data and specimen collection

Active, weekly ARI surveillance using a standard case definition was conducted seasonally from October to May in 2010-2014, and beginning in October 2014, year-round surveillance was performed. From 2010-2014 all respiratory specimens associated with an illness were collected by study personnel at illness visits. Starting in 2014, adult household members were trained to also collect nasal specimens from themselves and their participating children at study enrollment visits and were instructed to collect specimens upon onset of a respiratory illness in addition to specimens collected at study visits.^10^ Specimens collected at study visits were the default specimens tested; however, there were several instances in which self-collected specimens were used, such as when participants were unable to attend illness visits. After the spring of 2020, all respiratory specimens were self-collected. When a household reported an illness, study staff contacted the household to coordinate testing for those that met the ARI case definition. The case definition required two or more of the following symptoms: fever/feverishness, cough, nasal congestion, sore throat, body aches, chills, and headache; starting in 2014-2015, a separate ARI definition was used for those <3 years of age, requiring two or more of the following symptoms: fever/feverishness, cough, nasal congestion/runny nose, trouble breathing, fussiness/irritability, and decreased appetite. Study participants were not notified of their specimen testing results and were not explicitly instructed to isolate from household members. Longitudinal data on demographics, detailed health history, and influenza vaccination status were also collected at the annual enrollment/re-enrollment visits.

### Laboratory testing

Respiratory specimens were tested for influenza viruses using the U.S. Centers for Disease Control and Prevention (CDC) Influenza Virus Real-time reverse transcriptase polymerase chain reaction (RT-PCR) Influenza RUO Assays, then batch-tested for a panel of other respiratory viruses.^11^ Prior to the 2016-2017 study year, specimens were tested using singleplex reverse transcriptase polymerase chain reaction (RT-PCR) with primers and probes from the U.S. Centers for Disease Control and Prevention. Specimens collected during the 2016-2017 study season and beyond were tested using the Fast Track Diagnostics (FTD) Respiratory Pathogen 21 multiplex PCR kit (FTD-2-64-RUO, SMN: 11373928), Siemens Healthineers, Malvern, PA). Positive specimens were determined based on the presence of an amplification curve and virus-specific cycle threshold (Ct) was recorded for all positive results. Laboratory testing was performed at the University of Michigan School of Public Health.

### Study definitions

As study participants were required to meet a syndromic definition to be tested, the positive identification of a respiratory virus was considered an infection. Viral coinfection was defined as the simultaneous detection of more than one respiratory virus from a respiratory specimen. The household member who first exhibited symptoms prior to testing positive for a virus was considered the index case. If multiple household members developed symptoms on the same day, they were deemed co-index cases. A household transmission event occurred whenever a secondary case matching the respiratory pathogen of the index case within the same household occurred 1-14 days following the index case’s illness onset. The at-risk period for household members spanned the 14 days following a case’s illness onset and was extended each time another household member became a secondary case. The household’s illness cluster window ranged from the illness onset of the index case to 14 days following the final secondary case’s illness onset. When examining household transmission of each virus of interest, all initial viral infections within a household were preceded by at least 14 days without an identified infection caused by the virus of interest. A susceptible contact was determined to be infected with a virus other than the primary virus of interest if they tested positive for another virus during the household illness cluster window.

### Statistical analysis

We explored viral interaction by evaluating two main predictors of transmission: coinfection in the index case and detection of a virus other than the primary virus of interest in the susceptible contact. Household- and individual-level analyses were conducted three times—once for each virus of interest, including IAV, IBV, and RSV. For the IAV analysis, any non-IAV virus was considered as a coinfecting or co-circulating virus, including IBV and RSV. Similarly, any non-IBV virus was examined for interaction in the IBV analysis, and any non-RSV virus was examined in the RSV analysis.

To estimate the household risk of virus transmission within households with coinfected index cases while accounting for total time at risk, a household-level analysis was performed. Coinfection versus single infection among index cases was the primary predictor. Each household illness event was condensed into a single data point, and multivariable regression with Poisson mixed effects models was used to associate coinfection among index cases with the incidence of transmission of the virus of interest (IAV/IBV/RSV), adjusting for household size and age of the index case (<18 and ≥18 years). The outcome was an incidence rate ratio (IRR), defined as the ratio between the number of transmissions of the virus of interest divided by the total person-time at risk for those exposed to coinfected index cases versus those exposed to singly infected index cases. Random intercepts were included to account for household clustering. *P*-values were calculated using Wald tests at the 0.05 level.

To estimate an individual’s risk of infection when multiple viruses circulated simultaneously within their household while accounting for individual-level factors, an individual-level analysis was performed. The main predictor was a four-category variable with different combinations of exposure to a coinfected index case and infection with a virus other than the primary virus of interest (Figure 1). Category 1 represents individuals who were not infected with another virus and were exposed to a singly infected index case; category 2 includes those who were not infected with another virus and were exposed to a coinfected index case; category 3 includes those who were infected with another virus and exposed to a singly infected index case; category 4 represents those who were infected with another virus and were exposed to a coinfected index case. We examined the association between this predictor and the transmission of each virus of interest using mixed-effects logistic regression models with random intercepts to account for household clustering. The outcome was an odds ratio (OR), defined as the odds of having been exposed to the primary predictor among secondary cases with the virus of interest divided by the odds of having been exposed to the primary predictor among non-cases. The age of the index case (<18 and ≥18 years) was included as a household-level covariate. Individual-level covariates included sex, age group (0-5, 6-11, 12-17, 18-49, and 50+), and vaccination status (receipt of seasonal influenza vaccine ≥14 days prior to household exposure event) of susceptible contacts. *P*-values were calculated using Wald tests at the 0.05 level.

**Figure 1.**
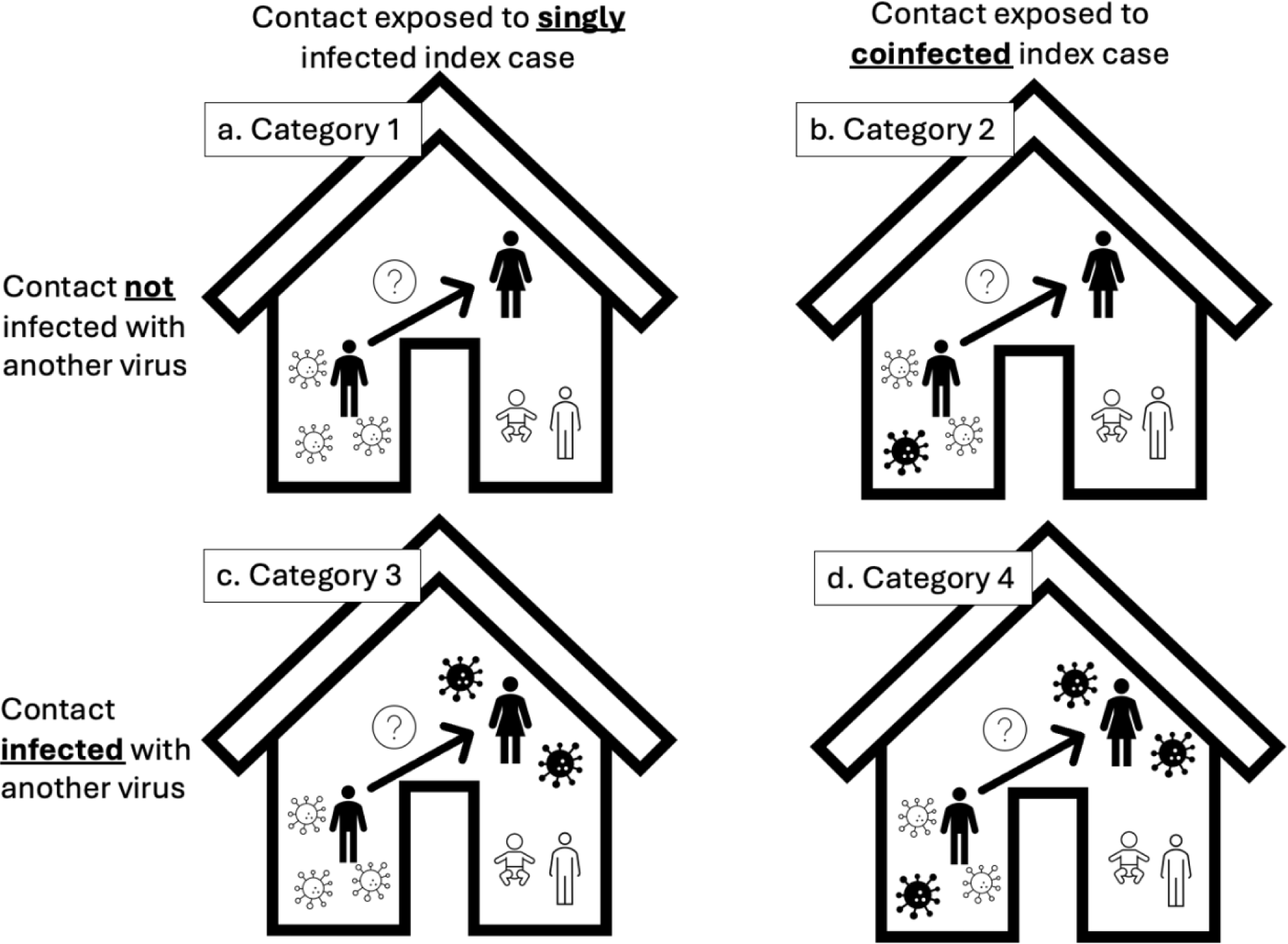
Four-category exposure variable used in multivariable mixed effects logistic regression models.

Covariates were selected a priori using a causal inference framework. Presence of at least one underlying high-risk comorbidity was considered for inclusion, but there was a small proportion of study participants with any high-risk comorbidity, so this variable was excluded from analyses. Sex, age group, and influenza vaccination status were compared by case status of each virus to the overall HIVE cohort study population from 2010-2020.

We have conducted three sensitivity analyses. First, the individual-level analysis was conducted using a transmission definition that required a positive test 2-14 days following the illness onset of the index case, rather than 1-14 days, to account for potential misclassification of co-index cases as secondary cases. This sensitivity analysis was performed for IAV and RSV models, but we were unable to do so for IBV due to sample size constraints. Next, as a proxy for viral load, another sensitivity analysis included the RT-PCR cycle threshold (Ct) value of the virus of interest for index cases as a continuous variable in the individual-level models. A third sensitivity analysis stratified the individual-level analyses by age groups to the smallest level of granularity that still allowed for model convergence. For IAV, age was stratified into 0-5 years, 6-17 years, and 18+ years. For RSV, age was stratified into groups 0-17 and 18+. The analysis could not be stratified by age for the IBV analysis.

## Results

### Study population

From 2010-2020, 957 households participated in HIVE, with 4,029 total participants. During this time, 546 household clusters involved IAV, with 786 total cases; 231 clusters involved IBV, with 301 total cases; 370 clusters involved RSV, with 493 total cases (Tables 1-3). The sex of index cases, secondary cases, and uninfected contacts generally followed the pattern of the larger HIVE cohort from 2018-2019 (52.2% female v. 47.8% male) except IAV, in which males (56.1%) represented a greater proportion of secondary cases than females. For IAV, the age groups that made up the highest proportion of index cases were ages 6-11 (30.1%) and 18-49 (30.7%); the largest proportion of secondary cases were in the 18-49 age group (36.8%), which aligns with the distribution of the overall HIVE cohort (39.0%). For IBV, the highest proportion of index cases were in the 6-11 age group (40.9%), while the highest proportion of secondary cases were in the 0-5 age group (37.2%), which is greater than the proportion of the overall HIVE cohort comprised of ages 0-5 (25.3%). For RSV, the highest proportion of index (49.4%) and secondary (41.7%) cases were in the 0-5 age group. For all three viruses of interest, over 60% of index cases, secondary cases, and non-cases received the seasonal influenza vaccine.

**Table 1.** Characteristics of HIVE study participants involved in IAV household illness events from 2010-2020.

| Household Illness Events, No. |  | 546 |  |  |  |
| --- | --- | --- | --- | --- | --- |
| Mean (Range) Household Members |  | 5 (3-9) |  |  |  |
| Cases, No. |  | 786 |  |  |  |
| Index, No. (%) |  | 574 (73.0%) |  |  |  |
| Secondary, No. (%) |  | 212 (27.0%) |  |  |  |
| Characteristic |  | Index Cases | Secondary Cases | Uninfected Contacts | HIVE Study ** |
| Sex, n (%) | F | 289 (50.4%) | 93 (43.9%) | 811 (51.5%) | 2078 (51.6%) |
|  | M | 285 (49.7%) | 119 (56.1%) | 765 (48.5%) | 1951 (48.4%) |
| Age Group, n (%) | 0-5 | 130 (22.7%) | 53 (25.0%) | 266 (16.9%) | 1035 (25.7%) |
|  | 6-11 | 173 (30.1%) | 47 (22.2%) | 380 (24.1%) | 759 (18.8%) |
|  | 12-17 | 75 (13.1%) | 25 (11.8%) | 232 (14.7%) | 410 (10.2%) |
|  | 18-49 | 176 (30.7%) | 78 (36.8%) | 635 (40.3%) | 1463 (36.3%) |
|  | 50+ | 20 (3.5%) | 9 (4.3%) | 63 (4.0%) | 115 (2.9%) |
| Influenza Vaccination, n (%) | No | 154 (31.2%) | 59 (32.2%) | 426 (32.1%) | 1154 (28.6%) |
|  | Yes | 340 (68.8%) | 124 (67.8%) | 902 (67.9%) | 2202 (54.7%) |
|  | Missing | 80 | 29 | 248 | 673 |
| Coinfected, n (%) | No | 416 (72.5%) | 169 (79.7%) | 1555 (98.7%) | - |
|  | Yes | 158 (27.5%) | 43 (20.3%) | 21 (1.3%)* |  |

Note: Participants may have appeared multiple times as index or secondary cases or uninfected contacts if involved in more than one household illness event.
\*Coinfected with non-influenza A viruses
\*\*Characteristics of HIVE study participants, including sex, age group at time of initial enrollment, and influenza vaccination status for that year

**Table 2.** Characteristics of HIVE study participants involved in IBV household illness events from 2010-2020.

| Household Illness Events, No. |  | 231 |  |  |  |
| --- | --- | --- | --- | --- | --- |
| Mean (Range) Household Members |  | 5 (3-9) |  |  |  |
| Cases, No. |  | 301 |  |  |  |
| Index, No. (%) |  | 242 (80.4%) |  |  |  |
| Secondary, No. (%) |  | 59 (19.6%) |  |  |  |
| Characteristic |  | Index Cases | Secondary Cases | Uninfected Contacts | HIVE Study** |
| Sex, n (%) | F | 117 (48.3%) | 31 (52.5%) | 362 (50.8%) | 2078 (51.6%) |
|  | M | 125 (51.7%) | 28 (46.5%) | 351 (49.2%) | 1951 (48.4%) |
| Age Group, n (%) | 0-5 | 56 (23.1%) | 22 (37.2%) | 92 (12.9%) | 1035 (25.7%) |
|  | 6-11 | 99 (40.9%) | 15 (25.4%) | 164 (23.0%) | 759 (18.8%) |
|  | 12-17 | 38 (15.7%) | 4 (6.8%) | 109 (15.3%) | 410 (10.2%) |
|  | 18-49 | 44 (18.2%) | 17 (28.8%) | 317 (44.5%) | 1463 (36.3%) |
|  | 50+ | 5 (2.1%) | 1 (1.7%) | 31 (4.4%) | 115 (2.9%) |
| Influenza Vaccination, n(%) | No | 76 (36.5%) | 15 (30.6%) | 221 (36.0%) | 1154 (28.6%) |
|  | Yes | 132 (63.5%) | 34 (69.4%) | 393 (64.0%) | 2202 (54.7%) |
|  | Missing | 34 | 10 | 99 | 673 |
| Coinfected, n (%) | No | 186 (76.9%) | 48 (81.4%) | 711 (99.7%) | - |
|  | Yes | 56 (23.1%) | 11 (18.6%) | 2 (0.3%)** |  |

Note: Participants may have appeared multiple times as index or secondary cases or uninfected contacts if involved in more than one household illness event.
\*Coinfected with non-influenza B viruses
\*\*Characteristics of HIVE study participants, including sex, age group at time of initial enrollment, and influenza vaccination status for that year

**Table 3.** Characteristics of HIVE study participants involved in RSV household illness events from 2010-2020.

| Household Illness Events, No. |  | 370 |  |  |  |
| --- | --- | --- | --- | --- | --- |
| Mean (Range) Household Members |  | 5 (3 -10) |  |  |  |
| Cases, No. |  | 493 |  |  |  |
| Index, No. (%) |  | 385 (78.1%) |  |  |  |
| Secondary, No. (%) |  | 108 (21.9%) |  |  |  |
| Characteristic |  | Index Cases | Secondary Cases | Uninfected Contacts | HIVE Study** |
| Sex, n (%) | F | 203 (52.7%) | 61 (56.5%) | 584 (50.9%) | 2078 (51.6%) |
|  | M | 182 (47.3%) | 47 (43.5%) | 564 (49.1%) | 1951 (48.4%) |
| Age Group, n (%) | 0-5 | 190 (49.4%) | 45 (41.7%) | 185 (16.1%) | 1035 (25.7%) |
|  | 6-11 | 82 (21.3%) | 23 (21.3%) | 256 (22.3%) | 759 (18.8%) |
|  | 12-17 | 34 (8.8%) | 6 (5.6%) | 134 (11.7%) | 410 (10.2%) |
|  | 18-49 | 70 (18.2%) | 31 (28.7%) | 534 (46.5%) | 1463 (36.3%) |
|  | 50+ | 9 (2.3%) | 3 (2.8%) | 39 (3.4%) | 115 (2.9%) |
| Influenza Vaccination, n(%) | No | 89 (25.8%) | 27 (24.5%) | 278 (27.9%) | 1154 (28.6%) |
|  | Yes | 256 (74.2%) | 79 (74.5%) | 717 (72.1%) | 2202 (54.7%) |
|  | Missing | 40 | 2 | 153 | 673 |
| Coinfected, n (%) | No | 235 (61.0%) | 77 (71.3%) | 1130 (98.4%) | - |
|  | Yes | 150 (39.0%) | 31 (28.7%) | 18 (1.6%)** |  |

Note: Participants may have appeared multiple times as index or secondary cases or uninfected contacts if involved in more than one household illness event.
\*Coinfected with non-RSV viruses
\*\*Characteristics of HIVE study participants, including sex, age group at time of initial enrollment, and influenza vaccination status for that year

### Household transmission

The proportion of secondary cases was higher for IAV (27.0%) compared to IBV (19.6%) and RSV (21.9%). The proportion of coinfected cases was higher for RSV compared to the influenza viruses, with 39.0% of index cases and 28.7% of secondary cases coinfected, compared to IAV (27.5% of index, 20.3% of secondary) and IBV (23.1% of index, 18.6% of secondary). The two most identified co-infecting and co-circulating viruses were rhinovirus/enterovirus and seasonal coronaviruses (Figure 2).

**Figure 2.**
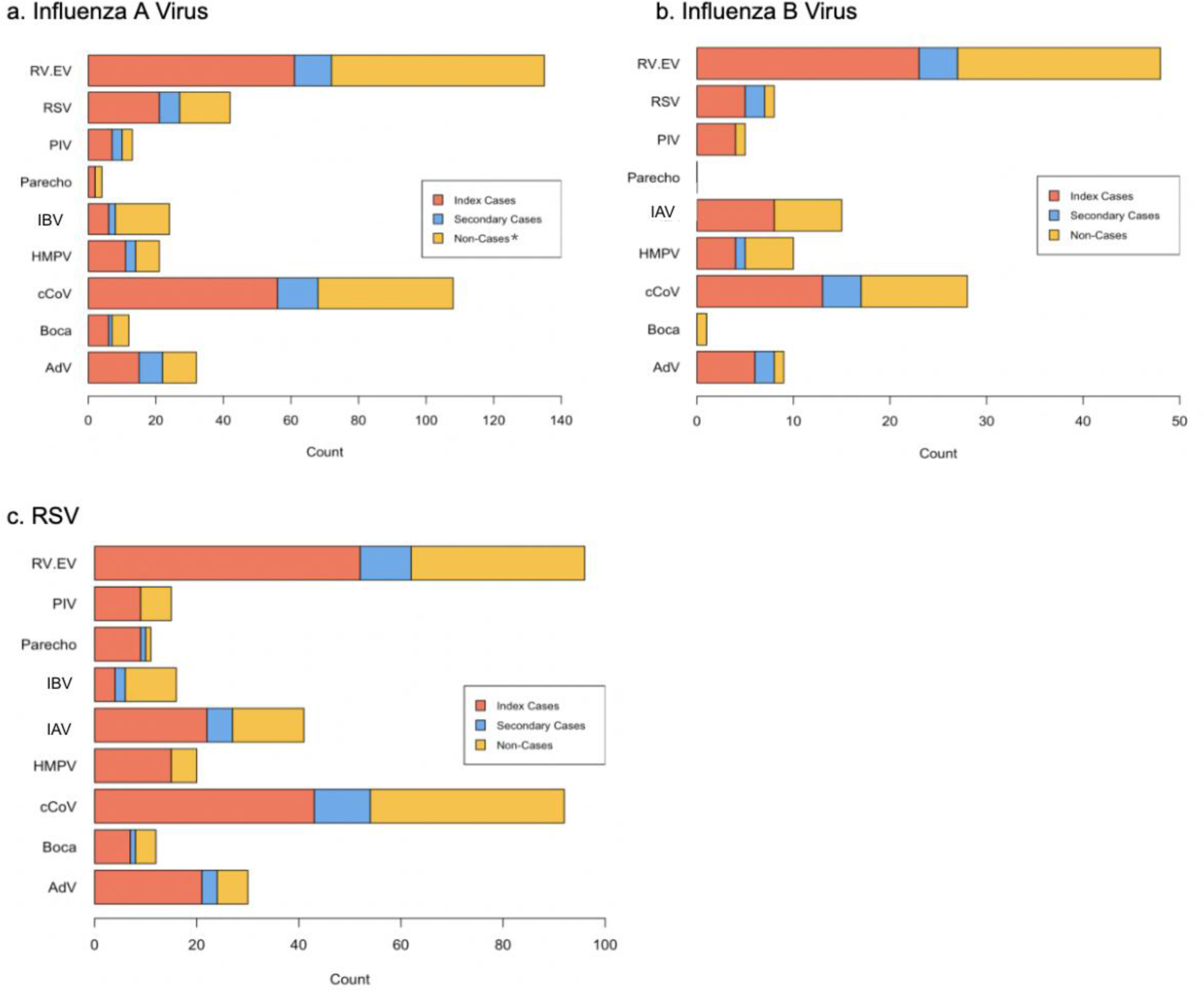
Viruses detected during household illness events, by primary virus of interest. AdV = adenovirus, Boca = bocavirus, cCoV = common/seasonal coronaviruses, HMPV = human metapneumovirus, IAV = influenza A virus, IBV = influenza B virus, Parecho = parechovirus, PIV = parainfluenza virus, RSV = respiratory syncytial virus, RV.EV = rhinovirus/enterovirus *Non-cases are those who were involved in a household illness cluster event for a virus of interest and did not test positive for the virus of interest.

### Household-level analyses

At the household-level, the presence of a coinfected index case in the household was associated with a lower risk of transmission to additional household members for IAV (IRR 0.44; 95% CI 0.29 – 0.66, *P*<0.001) and RSV (IRR 0.51; 95% CI 0.30 – 0.86, *P*=0.01) (Table 4). Having an index case <18 years of age was associated with a greater risk of transmission for IAV (IRR 2.13; 95% CI 1.46 – 3.10, *P*<0.001). The presence of a coinfected index case and an index case <18 years of age had no significant association with the risk of transmission of IBV. There was no significant association between the number of household members and the risk of transmission for any virus.

**Table 4.** Results from a multivariable regression using Poisson mixed effects models examining the association between exposure to coinfected index cases and the incidence of virus transmission.

| Characteristic | IRR (95% CI) | P-value |
| --- | --- | --- |
| IAV Model |  |  |
| Coinfecting index Case | 0.44 (0.29 – 0.66) | **<0.001** |
| No. Household Members | 0.91 (0.76 – 1.10) | 0.346 |
| Index case <18 | 2.13 (1.46 – 3.10) | **<0.001** |
| IBV Model |  |  |
| Coinfecting index Case | 0.85 (0.39 – 1.84) | 0.672 |
| No. Household Members | 1.29 (0.94 – 1.78) | 0.116 |
| Index case <18 | 1.07 (0.47 – 2.42) | 0.868 |
| RSV Model |  |  |
| Coinfecting index Case | 0.51 (0.30 – 0.86) | **0.01** |
| No. Household Members | 1.03 (0.81 – 1.32) | 0.80 |
| Index case <18 | 1.24 (0.64 – 2.42) | 0.53 |

### Individual-level analyses

At the individual level, exposure to a coinfected index case was associated with reduced transmission of IAV (OR 0.39; 95% CI 0.23-0.64, *P*<0.001) and RSV (OR 0.28; 95% CI 0.13-0.60, *P*=0.001) (Table 5). Infection with another virus among household contacts was associated with an increase in transmission of IAV (OR 3.49; 95% CI 2.02-6.03, *P*<0.001) and RSV (OR 6.05; 95% CI 2.55-14.35, *P*<0.001). Exposure to a coinfected index case among contacts infected with a different virus was associated with an increased transmission of IBV (OR 5.39; 95% CI 1.36-21.37, *P*=0.016), but this association was not seen for IAV or RSV. Male sex was positively associated with IAV transmission (OR 1.40; 95% CI 1.0.1-1.93, *P*=0.043), but not the other viruses. Exposure to an index case <18 years of age was positively associated with transmission of IAV (OR 2.58; 95% CI 1.70-3.93, *P*<0.001). Contacts under five years of age were more at risk for the transmission of all viruses of interest, except in the comparison between ages 0-5 and 50+ for IBV (OR 0.10; 95% CI 0.01-0.97, *P*=0.047). Receipt of the seasonal influenza vaccine ≥14 days prior to household exposure was associated with an increase in transmission of RSV (OR 2.06; 95% CI 1.11-3.81, *P*=0.021); there was no significant association for IAV or IBV.

**Table 5.**
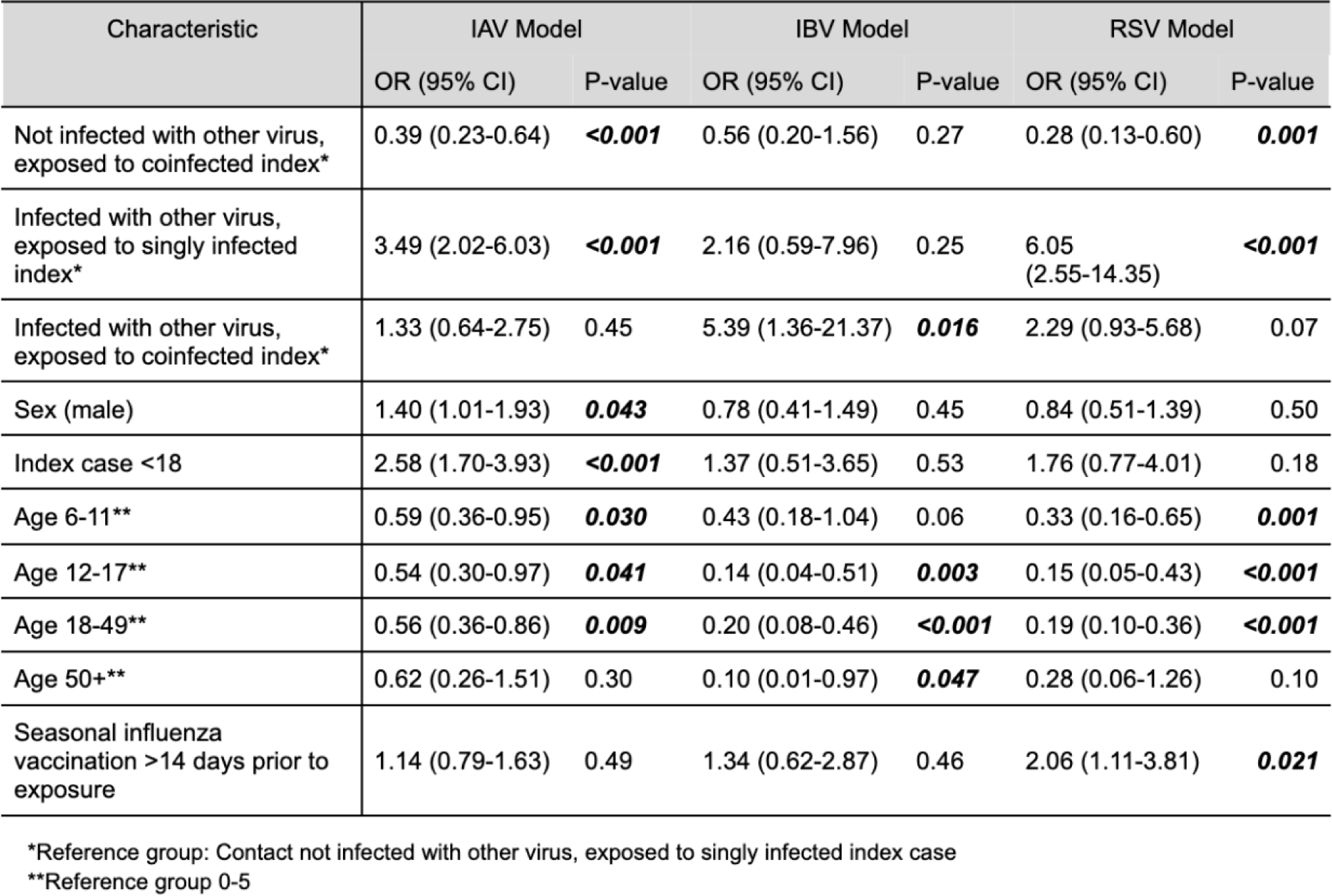
Results from multivariable mixed effects logistic regression models examining the association between infection with a different virus and exposure to a coinfected index case with the odds of virus transmission.

| Characteristic | IAV Model |  | IBV Model |  | RSV Model |  |
| --- | --- | --- | --- | --- | --- | --- |
|  | OR (95% CI) | P-value | OR (95% CI) | P-value | OR (95% CI) | P-value |
| Not infected with other virus, exposed to coinfecting index* | 0.39 (0.23-0.64) | **<0.001** | 0.56 (0.20-1.56) | 0.27 | 0.28 (0.13-0.60) | **0.001** |
| Infected with other virus, exposed to singly infected index* | 3.49 (2.02-6.03) | **<0.001** | 2.16 (0.59-7.96) | 0.25 | 6.05 (2.55-14.35) | **<0.001** |
| Infected with other virus, exposed to coinfecting index* | 1.33 (0.64-2.75) | 0.45 | 5.39 (1.36-21.37) | **0.016** | 2.29 (0.93-5.68) | 0.07 |
| Sex (male) | 1.40 (1.01-1.93) | **0.043** | 0.78 (0.41-1.49) | 0.45 | 0.84 (0.51-1.39) | 0.50 |
| Index case <18 | 2.58 (1.70-3.93) | **<0.001** | 1.37 (0.51-3.65) | 0.53 | 1.76 (0.77-4.01) | 0.18 |
| Age 6-11** | 0.59 (0.36-0.95) | **0.030** | 0.43 (0.18-1.04) | 0.06 | 0.33 (0.16-0.65) | **0.001** |
| Age 12-17** | 0.54 (0.30-0.97) | **0.041** | 0.14 (0.04-0.51) | **0.003** | 0.15 (0.05-0.43) | **<0.001** |
| Age 18-49** | 0.56 (0.36-0.86) | **0.009** | 0.20 (0.08-0.46) | **<0.001** | 0.19 (0.10-0.36) | **<0.001** |
| Age 50+** | 0.62 (0.26-1.51) | 0.30 | 0.10 (0.01-0.97) | **0.047** | 0.28 (0.06-1.26) | 0.10 |
| Seasonal influenza vaccination >14 days prior to exposure | 1.14 (0.79-1.63) | 0.49 | 1.34 (0.62-2.87) | 0.46 | 2.06 (1.11-3.81) | **0.021** |

\*Reference group: Contact not infected with other virus, exposed to singly infected index case
\*\*Reference group 0-5

### Sensitivity analyses

Changing the transmission timeframe from 1-14 days following the index case’s illness onset to 2-14 days resulted in 43 IAV cases, 16 IBV cases, and 19 RSV cases being reclassified from secondary to index; this did not substantially impact the direction or magnitude of the effect estimates (Supplemental Table 1). Similarly, the inclusion of Ct value for viruses of interest among index cases did not change the direction or magnitude of association (Supplemental Table 2). When the IAV model was stratified by age, exposure to a coinfected index case remained negatively associated with IAV transmission for all ages. When examining the relationship between contacts infected with another virus and transmission of IAV, stratifying by age resulted in different effect estimates for each age group, with an OR of 5.02 (95% CI 1.75-14.35, *P*=0.003) for those aged 0-5, 3.60 (95% CI 1.35-9.58, *P*=0.010) for those aged 6-17, and 2.43 (95% CI 0.80-7.42, *P*=0.118) for those aged 18+. Exposure to a coinfected index case among contacts infected with a different virus was associated with reduced transmission for those aged 0-5 (OR 0.60; 95% CI 0.11-3.42, *P*=0.569) and 18+ (OR 0.86; 95% CI 0.21-3.54, *P*=0.835), but was associated with an increased transmission for those aged 6-17 (OR 3.54; 95% CI 1.17-10.66, *P*=0.025). For RSV, exposure to a coinfected index case remained negatively associated with transmission for all age strata; infection with another virus among contacts remained positively associated with transmission for all age strata. There was not a strong relationship between RSV transmission and exposure to a coinfected index case among contacts infected with a different virus for any age.

## Discussion

We used data from a prospective cohort study of households with children in Southeast Michigan to examine the effects of viral interaction on household transmission of respiratory viruses. Separate analyses were conducted for three different viruses of interest: IAV, IBV, and RSV. Coinfection among index cases was associated with a reduced transmission of IAV and RSV in individual-level and household-level models. Adjusting for Ct value of viruses of interest among index cases in the individual-level models did not change this relationship, indicating that the Ct value of the index case alone does not explain the protective association seen when participants were exposed to coinfected index cases. Infection with a different virus among household contacts was associated with increased transmission of IAV and RSV. The observed relationships between the main predictors and virus transmission remained when the models were stratified by age; however, when examining the relationship between infection with another virus and acquisition of IAV, stratifying by age resulted in different effect estimates for each age group, indicating that age could be a potential effect modifier in this relationship.

The possibility of viral interaction affecting virus transmission patterns has been hypothesized but primarily has been observed in ecologic studies that are unable to directly evaluate the mechanisms behind this phenomenon. Casalegno et al. (2010) evaluated the hypothesis that rhinoviruses delayed the onset of the A(H1N1) 2009 influenza virus pandemic in France.^12^ The authors found that between weeks 36 and 48 of 2009, both rhinoviruses and H1N1 were detected but in different timeframes. During a three-week cocirculation period of these two viruses, rhinovirus detection appeared to reduce the likelihood of H1N1 detection, supporting the hypothesis that rhinovirus infection can inhibit H1N1 infection. Another study by van Asten et al. (2016) investigated time trends and correlation between eight common viruses in the Netherlands over a ten-year period.^13^ The authors found that when IAV epidemics occurred earlier than usual, the epidemics of three other viruses were affected; RSV waves tended to be delayed, coronavirus outbreaks were intensified, and IBV tended to not appear at all.

Virus-virus interaction may manifest in several ways at the cellular, host, and population levels. Examples of cellular-level interaction may involve competition for host resources or certain viruses enhancing or inhibiting replication of other viruses.^6^ At the host-level, viruses may work synergistically or antagonistically to result in differences in disease severity than would occur if infected with only one of the viruses.^14^ At the population-level, viral interaction may result in more or less frequent coinfection of certain virus combinations than would be expected by chance alone, or may impact the circulation of multiple viruses within a population.^15,16^ The type of virus-virus interaction observed may depend on the specific combination of viruses involved.^7^ Studies of viral interaction have historically been conducted at the host level through animal models, using clinical and cross-sectional data to assess illness severity, and ecologically to quantify interference at the population level by examining the timing of virus waves. ^15,17–21^ We are aware of only one study that has examined the relationship between viral coinfection and virus transmission in humans. This study by Scott et al. (2019) considered various predictors of household virus transmission in rural Nepal, and the authors found that viral coinfection among index cases was a risk factor for household transmission.^22^ Possible reasons for the differing findings between this study and the current study include differences in study populations, viruses tested, and ARI case definitions.

Our findings suggest that respiratory virus transmission may be impacted by other viruses co-infecting within individuals and co-circulating within households. Rhinovirus/enterovirus and coronaviruses were the most commonly identified co-infecting and co-circulating viruses. Rhinovirus has been shown to interfere with RSV and IAV infections.^23,24^ Pinky and Dobrovolny (2016) used a mathematical model to investigate the kinetics of viral coinfections within the respiratory tract.^25^ They found that while rhinovirus was seemingly unaffected by the presence of other viruses, the replication of other viruses was suppressed in the presence of rhinovirus. As for the current study, one hypothesis is that the observed protective effect of exposure to coinfected index cases may be driven by this antagonistic relationship between rhinovirus and RSV/IAV, which could inhibit the transmission of these viruses of interest. Kim et al. (2024) examined coinfections between influenza viruses and human OC43 coronavirus in normal human bronchial epithelial cells and found that while coinfection with OC43 did not affect replication of IAV or IBV, select cytokine/chemokine expression was increased in coinfected cells compared to singly infected cells.^26^ A hypothesis for our other main finding is that coinfection among secondary cases with IAV or RSV plus another virus led to more symptomatic illness than infection with either of the viruses alone; because the HIVE study requires a standard ARI case definition for testing, these virus combinations would be detected more frequently if they more often resulted in illnesses that met the ARI definition. Studies comparing disease severity between coinfections and single virus infections have had mixed results.^27^ However, these studies typically use data from clinical settings with cases that require medical care and/or hospitalization. With a wider range of illness severity observed in the community setting, it is possible that coinfections involving IAV or RSV result in more symptomatic illness than single infections. Regardless, our findings suggest that it may be important to consider interaction within the index case and among susceptible contacts when evaluating the transmission implications of viral interaction.

The age of household members may impact household transmission dynamics for many reasons. For example, Munywoki et al. (2014) studied RSV illnesses within households in Kenya and found that school-aged children frequently were index cases within households, often leading to transmission to infant siblings.^28^ Also, having received a seasonal influenza vaccination at least 14 days prior to household exposure was not associated with reduced transmission of IAV or IBV and was associated with an increased transmission of RSV. Cowling et al. (2012) identified an increased risk of non-influenza viruses after receipt of an inactivated influenza vaccine, and they proposed that this finding was related to a lack of non-specific immunity against other respiratory viruses in the absence of influenza infection.^29^ It is possible that this phenomenon is responsible for the apparent risk of RSV infection associated with receipt of influenza vaccination observed in the present study. Notably, the HIVE population may not be representative of many other populations, as it has relatively high levels of influenza vaccination. While not the focus of the present study, a more comprehensive analysis focused on influenza vaccination and the risk of non-influenza viruses could disentangle this relationship.

This study has multiple key strengths. To our knowledge, this is the first study that explicitly examines virus-virus associations in the context of virus transmission. The use of a prospective cohort study allowed for households to be followed for the entirety of their illness cluster windows. The breadth of data available through the HIVE study over the ten-year period from 2010-2020 allowed us to use a multivariable analysis to examine multiple potential modes of virus-virus interaction for three different viruses of interest. Because HIVE is a community-based study that utilizes active ARI surveillance to identify illnesses, less severe illnesses that did not require medical attention—and therefore are often excluded from studies that utilize clinical data—were captured. The inclusion of mild illnesses is crucial for the comprehensive understanding of virus transmission, especially when virus-virus interaction may be involved. Also, studying virus transmission within the household setting is ideal, as household exposures to respiratory viruses represent the highest-risk exposures, often involving interactions with close proximity for long durations of time.

Our study has several limitations. The detection of a virus in the respiratory tract does not necessarily indicate a current infection, but rather may indicate a recently resolved infection or asymptomatic shedding. Similarly, the co-detection of viruses may not represent true coinfection; however, viral interaction may be present when one virus infects a host subsequently after another.^23^ Therefore, despite the inability to differentiate between concurrent and successive infections, viral interaction could have occurred, nonetheless. In the absence of serially testing study participants, it is not possible to establish temporality of viral infections. This could have resulted in misclassification of some index and secondary cases as coinfected when they did not truly experience concurrent infection with both viruses. However, the absence of concurrent infections does not exclude the possibility of viral interaction that may have occurred as a result of consecutive infections with different viruses. Also, we were likely unable to completely account for underlying differences in susceptibility that may be common for the acquisition of multiple viruses, which is often a limitation in studies of ARI. Finally, there was the possibility of misclassification of secondary cases in the absence of sequencing to confirm household transmission via matching viral strains. However, the high-risk nature of household exposures suggests that this misclassification may be minimal.

Understanding the ways in which influenza viruses and RSV interact with other viruses could inform public health planning and prevention efforts, especially in preparation for seasons with exceptionally high levels of these viruses or the circulation of multiple viruses simultaneously, as transmission may be heightened or reduced depending on the viruses involved. As Opatowski et al. (2018) concluded, evaluating influenza viruses in isolation may lead to an incomplete picture of the burden influenza viruses pose, which in turn may hinder public health prevention efforts.^30^ The same could be argued for RSV. It is important for more studies to examine the transmission implications of virus-virus interaction in order to get a more complete understanding of the population impact of viral interaction.

## Conclusion

This study provides novel insight into the ways in which virus-virus interaction may impact the transmission of IAV, IBV, and RSV within households. This interaction may occur at the level of the individual introducing the virus into the household, blocking transmission to susceptible household members. This interaction also may occur within the susceptible contact, encouraging dual infection of certain viruses. Future research should focus on delving further into the interaction dynamics of different virus combinations, as well as utilizing serial testing to identify asymptomatic infections and establish infection temporality.

## Data Availability

All data produced in the present study are available upon reasonable request to the authors.

## Acknowledgements

The authors would like to thank the HIVE study participants for their participation and the HIVE study staff for their dedication to this project. The HIVE study was supported by funding from the Centers for Disease Control, the National Institute of Allergy and Infectious Disease and the National Institutes of Health (75N93021C00015, HHSN272201400005C, U01IP001034, R56 AI097150, R01AI097150-01A1, U01 IP000474, U18IP000170). This work was also supported by funding for the MHome study through the National Institute of Allergy and Infectious Diseases, under R01 AI148371.

## Contributions

JCI conceptualized the project, planned and performed the statistical analysis, interpreted the results, and drafted and revised the article. ETG validated the statistical analysis and revised the article. APC coordinated the HIVE study and revised the article. MRS and APC performed data curation. EJ performed HIVE specimen testing. RT oversaw and performed HIVE specimen testing and revised the article. MCE, ASL, SC and ASM provided intellectual guidance and revised the article. ETM conceptualized and oversaw all aspects of the project and revised the article.

## Corresponding author

Correspondence to Jessica C. Ibiebele.

## Ethics Declarations

The authors declare no competing interests.

