## Supplemental Tables 1-4 for "The impacts of viral interaction on household transmission of respiratory viruses"

**Supplementary Table 1.** Sensitivity analysis using a different transmission definition of 2-14 days following illness onset of index cases for IAV and RSV

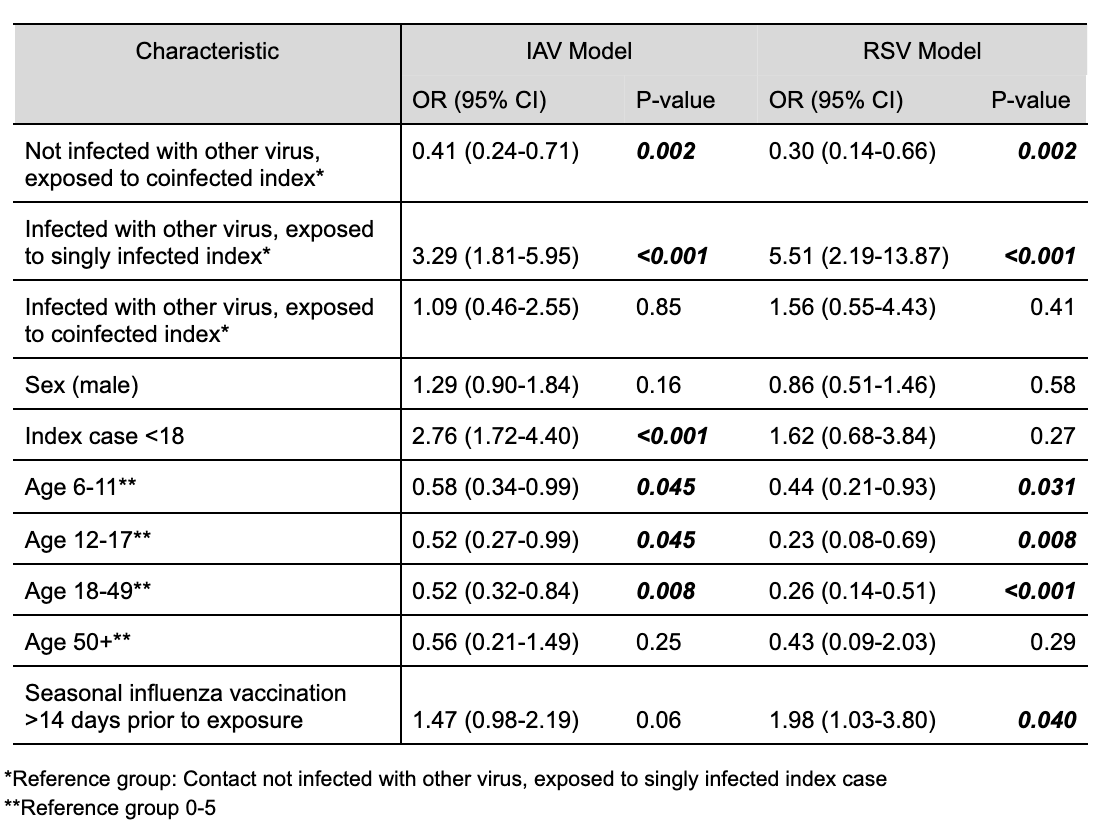

**Supplementary Table 2.** Sensitivity analysis including additional covariate representing Ct value of the virus of interest for index cases

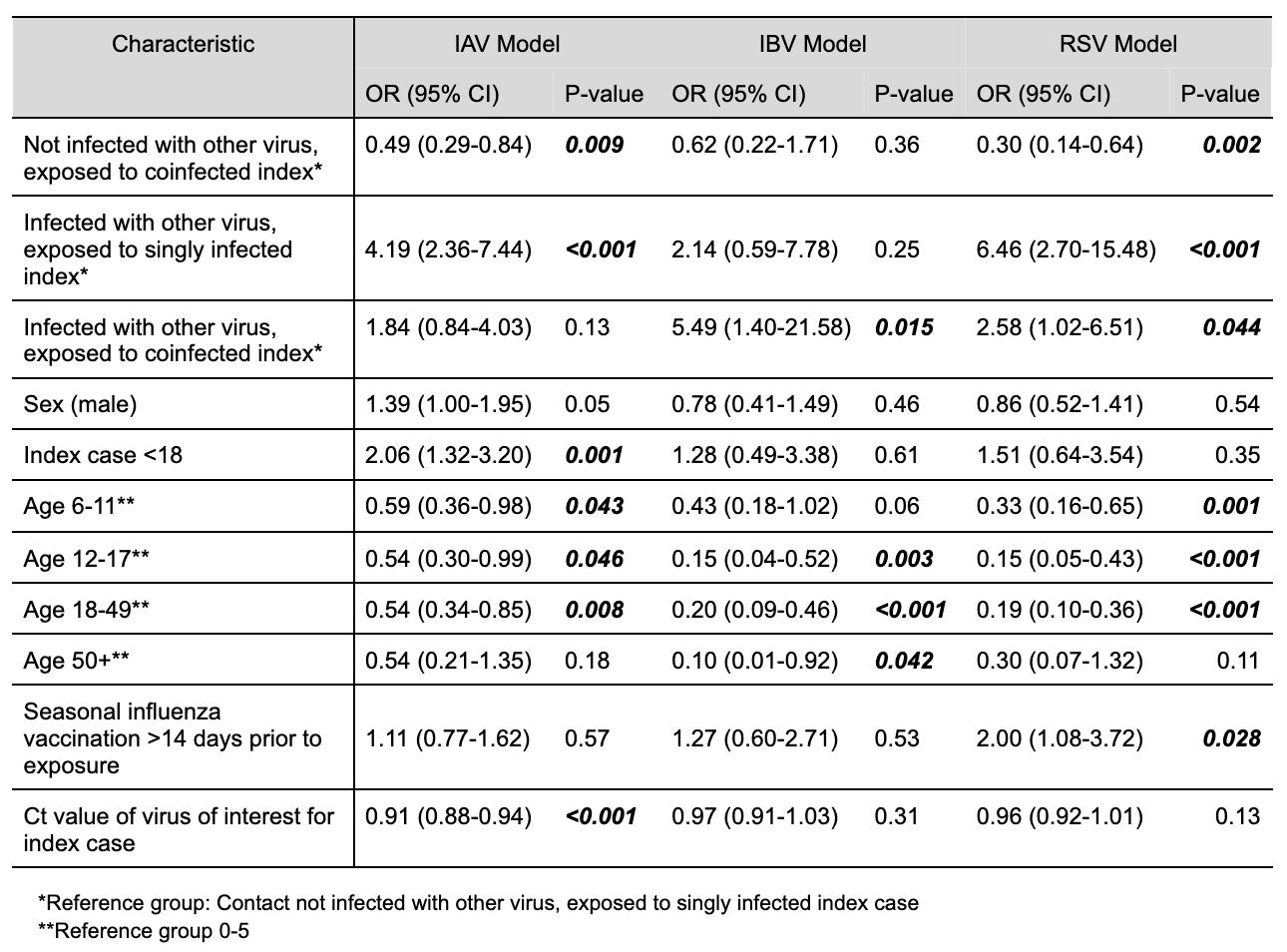

**Supplementary Table 3.** Sensitivity analysis stratifying by age groups 0-5, 6-17, and 18+ for IAV household transmission events

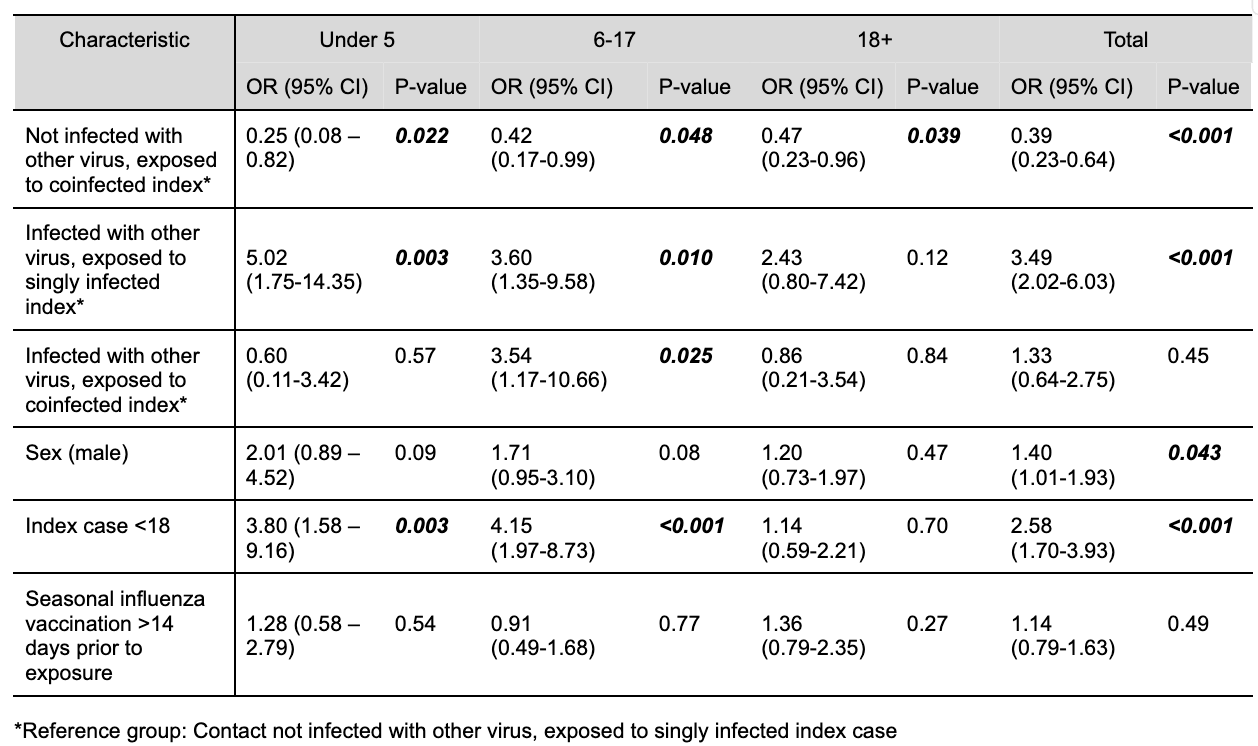

**Supplementary Table 4.** Sensitivity analysis stratifying by age groups 0-17 and 18+ for RSV household transmission events

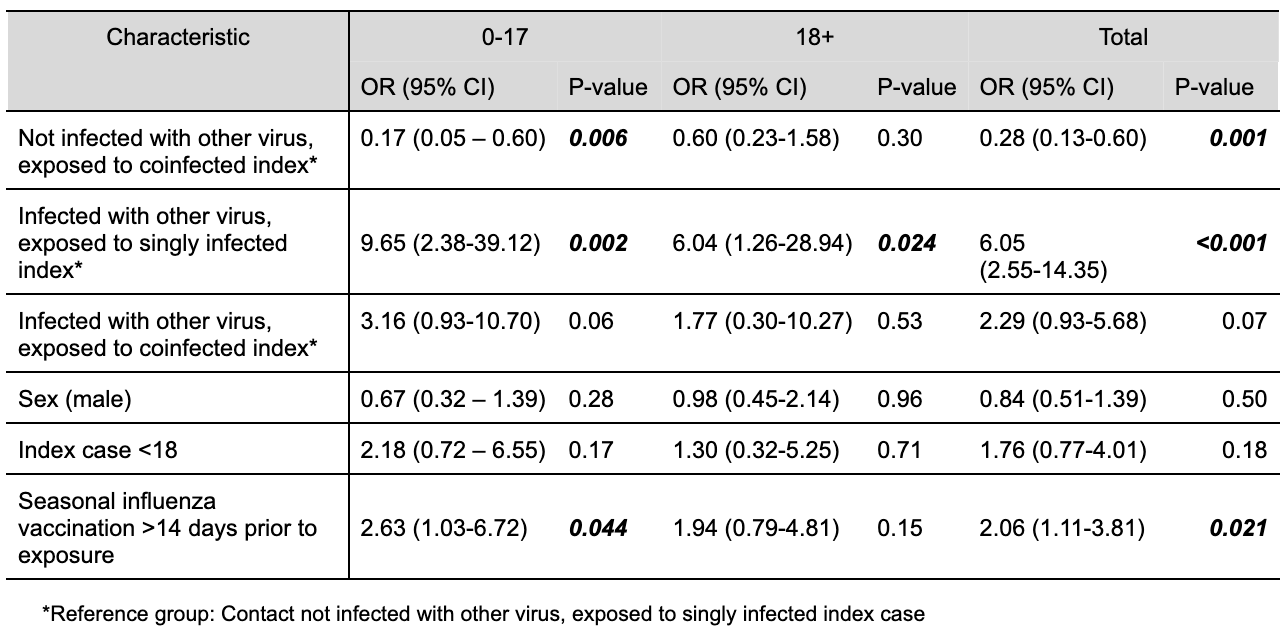
